# Minimal Detectable Change in Gait Biomechanics Post-Stroke: Disentangling the Effects of Walking Speed and Stroke-Related Variability

**DOI:** 10.64898/2026.08.25.26361347

**Authors:** Alejandro Aguirre Ramirez, Andrian Kuch, Russell T. Johnson, Natalia Sanchez

## Abstract

Impaired motor control post-stroke results in reduced walking speeds and increased gait variability. This variability reduces reliability and makes identifying longitudinal changes via gait analysis difficult since changes may occur within the margin of measurement error. We quantified intra-class correlation coefficients (ICC) and minimal detectable change (MDC) in post-stroke individuals and neurotypical individuals walking at matched speeds, to isolate the impact of gait speed and post-stroke impairments on gait-analysis reliability.

We collected gait data over two days from N=15 post-stroke individuals walking on a treadmill at their self-selected speed, and from N=13 age- and sex-matched neurotypical controls walking at both their self-selected speed and a speed matched to a post-stroke participant. We calculated ICC and MDC values for spatiotemporal variables, bilateral joint ranges of motion (ROM), and bilateral peak propulsive and peak vertical ground reaction forces (GRF).

Spatiotemporal ICCs showed excellent reliability across groups (range [0.813-0.988]), yet MDC values were greater post-stroke than in speed-matched controls. ICCs for joint ROM ranged from poor to excellent reliability across groups ([0.362-0.960]). Post-stroke joint ROM MDCs were 27%-53% of the gait ROM compared to 11%-42% in neurotypical controls. ROM MDCs were greater in the non-paretic compared to the paretic extremity. ICC for peak GRFs showed good to excellent reliability across groups (range [0.778-0.980]), with post-stroke peak GRF MDCs greater than in speed-matched controls.

Our results suggest that stroke-related neuromotor impairments influence reliability beyond the effects of walking speed alone, and we provide quantitative MDC benchmarks for interpreting gait changes post-stroke following clinical interventions.

## Introduction

Gait analyses are widely used to quantify locomotor impairments, evaluate rehabilitation outcomes, and investigate the biomechanics underlying pathological gait (Baker, 2006; Kadaba et al., 1990; Perry and Burnfield, 2010). To derive meaningful clinical and scientific insights from gait analyses, changes observed following interventions, recovery, or disease progression must reflect true changes in motor performance rather than normal behavioral variability or low measurement reliability due to errors (Portney, 2020). This distinction is important in pathological populations, where gait patterns are often characterized by substantial variability (Hausdorff, 2005; Kiss, 2011; Winter, 1984). Consequently, interpreting longitudinal changes in gait remains challenging because observed differences may reflect the biomechanical adaptations related to the phenomenon under investigation, natural variability in motor behaviors (Winter, 1984), methodological inconsistencies inherent to gait analysis (Wolf et al., 2009), or, most likely, a combination of these factors.

Specifically, individuals post-stroke commonly exhibit increased variability in spatiotemporal gait characteristics due to impairments in motor control, balance, and interlimb coordination (Balasubramanian et al., 2009). Variability in gait spatiotemporal parameters post-stroke is often greater than in neurotypical individuals (Patel et al., 2022) and increases with impairment severity (Balasubramanian et al., 2009; Patel et al., 2022; Sanchez et al., 2021). While this variability reflects impaired neuromotor control post-stroke (Olney and Richards, 1996), it complicates interpreting longitudinal gait assessments because repeated measurements may vary independently of rehabilitation-induced improvements, recovery processes or injury mechanisms under study. The intraclass correlation coefficient (ICC) assesses the extent to which measurements can be replicated (Koo and Li, 2016; McGraw and Wong, 1996). The minimal detectable change (MDC) provides a statistical threshold representing the smallest change that can be interpreted as exceeding measurement error (Portney, 2020). Together, the ICC and MDC can help establish measurement reliability and are essential for determining whether observed changes in gait outcomes reflect true changes in performance in individuals post-stroke, rather than low reliability of gait analyses.

Previous research has reported ICC and MDC values for spatiotemporal and kinematic gait measures in individuals post-stroke during both overground and treadmill walking (Geiger et al., 2019; Kesar et al., 2011). A primary limitation of previous studies is that they did not compare post-stroke MDC values with those of neurotypical adults without neurological or musculoskeletal impairments, which would allow differentiation between methodological and pathological variability. Since walking speed is a major determinant of gait variability (Jordan et al., 2007; Kang and Dingwell, 2008), and individuals post-stroke typically walk slower than neurotypical adults (Olney and Richards, 1996; Sánchez et al., 2026), it is possible that results of prior work demonstrating greater MDCs for gait variables in the post-stroke population are due to the slower walking speed post-stroke. Comparing individuals post-stroke with age-, sex-, and speed-matched neurotypical controls will help determine whether gait variability, and consequently the reliability of gait analysis, differ in people post-stroke even when the effect of walking speed is accounted for.

The purpose of this study was to quantify ICC and MDC values for commonly reported gait outcomes in individuals post-stroke compared to speed-, age- and sex-matched controls. We assessed the ICC and MDC of kinematics, including spatiotemporal parameters and hip, knee, and ankle joint ranges of motion (ROM), which are frequent targets of post-stroke rehabilitation interventions (Lewek et al., 2018a; Park et al., 2021; Sánchez and Finley, 2018; Spencer et al., 2021; Teasell et al., 2003; Wonsetler and Bowden, 2017a). Given the established importance of paretic propulsion and limb loading as rehabilitation targets following stroke (Awad et al., 2015; Choe et al., 2024; Hsiao et al., 2020; Kim and Eng, 2004; Lewek et al., 2018b; Roelker et al., 2019; Shen et al., 2022; Swaminathan et al., 2023; Wonsetler and Bowden, 2017b) we also quantified the reliability of peak propulsive and peak vertical ground reaction forces (GRF). We hypothesized that ICC values would be lower and MDC values would be greater in individuals post-stroke than controls, reflecting added variability associated with stroke-related motor impairments that compound the effects of walking speed or methodological errors (Wolf et al., 2009) on gait analysis reliability. Establishing ICC and MDC thresholds for post-stroke gait relative to neurotypical gait will improve interpretation of intervention outcomes by distinguishing true biomechanical change from typical sources of measurement error in gait analysis.

## Methods

### Participants

Institutional Review Board of Chapman University (IRB #23-57 for participants post-stroke; IRB #23-12 for control participants) gave ethical approval for this work, and all participants provided written informed consent prior to data collection. Participants post-stroke were eligible if they: (1) had experienced a unilateral cerebrovascular accident more than six months prior to enrollment; (2) had no additional neurological or musculoskeletal conditions that could affect gait; (3) were able to walk continuously on a treadmill for at least 5 minutes without physical assistance (e.g., without a cane or walker) but, use of an ankle–foot orthosis or brace was permitted; and (5) were able to provide informed consent. Neurotypical control participants were included if they: (1) had no history of neurological or musculoskeletal conditions affecting gait; (2) were matched to a participant post-stroke by age and sex; (3) were able to walk continuously on a treadmill for at least 5 minutes without assistive devices; and (4) were able to provide informed consent.

### Experimental Protocol

Following informed consent, all participants underwent a series of clinical assessments. In participants post-stroke, lower-limb motor impairment was quantified using the lower extremity section of the Fugl-Meyer Assessment (FMA) (Fugl-Meyer, 1976), and the Functional Gait Assessment (Wrisley and Kumar, 2010) and balance was assessed using the Berg Balance Scale (BBS) (Berg, 1989). Self-selected walking speed (SSS) was evaluated in both groups using the 6-minute walk test (6MWT) (Fulk and He, 2018), and we calculated the average walking speed over the duration of the test.

Gait data were collected during treadmill walking. All participants completed two testing sessions separated by 1–3 days. Participants wore an overhead harness to prevent falls, without providing any body weight support. Control participants walked for 2 minutes at their SSS determined from the 6MWT and at a speed matched to that of an age- and sex-matched participant post-stroke (“Matched” condition). The Matched condition enabled comparisons between groups at slower walking speeds associated with increased variability (Jordan et al., 2007; Kang and Dingwell, 2008). Participants post-stroke walked for 2 minutes at their SSS. All participants walked without holding onto treadmill handrails.

### Data Acquisition

We collected kinematic and kinetic data using a Gait Real-time Analysis Interactive Lab (GRAIL; Motek Medical, Netherlands), equipped with a 10-camera motion capture system (sampling at 100 Hz) and an instrumented dual-belt treadmill (sampling at 1000 Hz). A total of 26 reflective markers were placed on anatomical landmarks according to a modified Human Body Model 2 to represent the trunk and lower limbs (Van Den Bogert et al., 2013).

### Data Processing and Analyses

We used the Gait Offline Analysis Tool (Motek Medical, Netherlands) to compute stride-by-stride spatiotemporal gait measures, joint kinematics in the sagittal plane, and ground reaction forces in the anterior-posterior and vertical directions. We used a low-pass filter with a cutoff frequency of 6 Hz to filter marker data. Heel strikes to identify individual strides were detected based on Zeni’s marker-based algorithm (Zeni et al., 2008). We obtained stride length, stride time, step width, and cadence. We obtained step length, as well as stance and swing times bilaterally. For each participant, we computed the average across all strides at the target SSS within each day. Peak propulsive and vertical GRFs (normalized to body weight = mass*9.81m/s^2^) and ranges of motion, defined as the difference between the maximum and minimum joint kinematics were extracted for further statistical analyses. Data were further analyzed using custom scripts written in MATLAB 2024a (MathWorks Inc., Natick MA).

### Statistical Analyses

Data were tested for normality using the Shapiro-Wilk test. For normally distributed data, we used independent-samples t-tests to compare data between post-stroke participants and controls. For non-normally distributed data, we used the Wilcoxon Rank-Sum test. To ensure that our samples were age-matched, we compared ages between post-stroke participants and controls using independent-samples tests. We compared spatiotemporal characteristics, range of motion, and peak propulsive and vertical GRFs between post-stroke participants and controls using independent-samples tests. All p-values were corrected for multiple tests using the Bonferroni correction.

We assessed test–retest reliability using the absolute agreement ICC (Salarian, 2026) calculated with a two-way random-effects model (Weir, 2005). We set the thresholds for reliability as in previous literature such that *ICC* < 0.50 indicate poor reliability, 0.5 ≤ *ICC* < 0.75 indicate moderate reliability, 0.75 ≤ *ICC* < 0.9 indicate good reliability and 0.9 ≤ *ICC* indicate excellent reliability (Koo and Li, 2016; Portney, 2020; Weir, 2005). Statistical significance for ICCs was set at p < 0.05 after correcting for multiple tests using a Bonferroni correction. From the ICC, we calculated the standard error of measurement 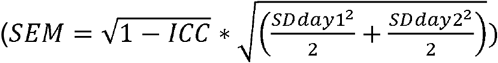, which we then used to obtain the 95% confidence interval MDC 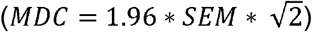.

## Results

N=15 participants post-stroke (51.4 ± 14.8 years; 8 female, 7 male; 9 left and 6 right paresis) and N=13 age-matched control participants (46.9 ± 12.24 years; 7 female, 6 male) were tested over two sessions. Participant demographics are summarized in Table 1.

**Table 1:** Descriptive Statistics.

|  | Stroke (N=15) | Control (N=13) |
| --- | --- | --- |
| Sex | 8F/7M | 7F/6M |
| Age (years) | 51.4 ± 14.8 | 46.9 ± 12.2 |
| Mass (kg) | 84.3 ± 14.5* | 65.8 ± 10.9 |
| Speed (SSS; m/s) | 0.71 ± 0.27* | 1.18 ± 0.15 |
| Time post-stroke (months) | 87.3 ± 65.8 |  |
| FMA | 23.1 ± 5.9 |  |
| BBS | 50.9 ± 4.5 |  |
| FGA | 17.3 ± 4.9 |  |
| Paresis | 9L/6R |  |

Data are reported when applicable as mean (SD)
\*p<0.001 significant differences between participants post-stroke and neurotypical controls
SSS: Self-Selected Speed

SSS was significantly faster in control participants (1.18 ± 0.15 m/s) compared to participants post-stroke (0.71 ± 0.27 m/s; *p* < 0.001). In contrast, during the Matched condition, walking speed did not differ between groups (controls: 0.86 ± 0.26 m/s; *p* = 0.154). At Matched speeds, no significant between-group differences were observed in any spatiotemporal parameters for either limb (*p* > 0.086).

No significant differences were observed between Day 1 and Day 2 for any variable; therefore, Tables 2-4 report mean values across days, with standard deviations reflecting between-subject variability in these averaged measures. No significant differences were observed between dominant and non-dominant limbs in control participants. Accordingly, we compared all variables between the non-dominant limb of controls and the paretic limb of participants post-stroke, and between the dominant limb of controls and the non-paretic limb of participants post-stroke.

**Table 2:**
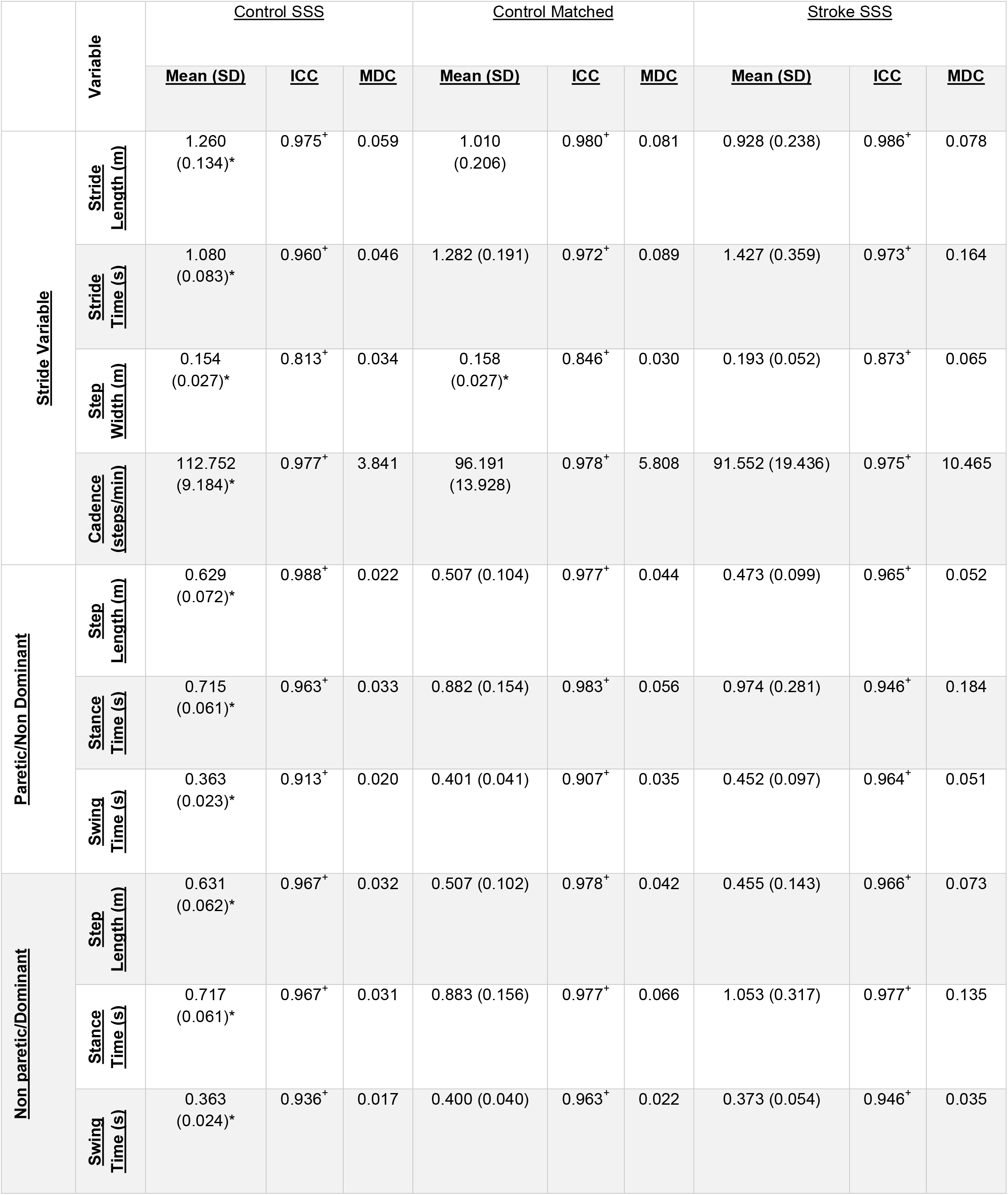
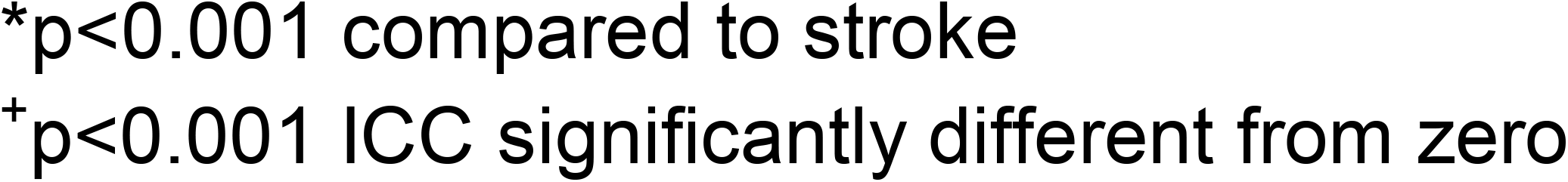
Mean (Standard Deviation), ICC, and MDC values for spatiotemporal variables for control participants walking at self-selected speed, controls walking at matched speed, and stroke participants.

|  | Variable | <u>Control SSS</u> |  |  | <u>Control Matched</u> |  |  | <u>Stroke SSS</u> |  |  |
| --- | --- | --- | --- | --- | --- | --- | --- | --- | --- | --- |
|  |  | <u>Mean (SD)</u> | <u>ICC</u> | <u>MDC</u> | <u>Mean (SD)</u> | <u>ICC</u> | <u>MDC</u> | <u>Mean (SD)</u> | <u>ICC</u> | <u>MDC</u> |
| <u>Stride Variable</u> | <u>Stride Length (m)</u> | 1.260<br>(0.134)* | 0.975<sup>+</sup> | 0.059 | 1.010<br>(0.206) | 0.980<sup>+</sup> | 0.081 | 0.928 (0.238) | 0.986<sup>+</sup> | 0.078 |
|  | <u>Stride Time (s)</u> | 1.080<br>(0.083)* | 0.960<sup>+</sup> | 0.046 | 1.282 (0.191) | 0.972<sup>+</sup> | 0.089 | 1.427 (0.359) | 0.973<sup>+</sup> | 0.164 |
|  | <u>Step Width (m)</u> | 0.154<br>(0.027)* | 0.813<sup>+</sup> | 0.034 | 0.158<br>(0.027)* | 0.846<sup>+</sup> | 0.030 | 0.193 (0.052) | 0.873<sup>+</sup> | 0.065 |
|  | <u>Cadence (steps/min)</u> | 112.752<br>(9.184)* | 0.977<sup>+</sup> | 3.841 | 96.191<br>(13.928) | 0.978<sup>+</sup> | 5.808 | 91.552 (19.436) | 0.975<sup>+</sup> | 10.465 |
| <u>Paretic/Non Dominant</u> | <u>Step Length (m)</u> | 0.629<br>(0.072)* | 0.988<sup>+</sup> | 0.022 | 0.507 (0.104) | 0.977<sup>+</sup> | 0.044 | 0.473 (0.099) | 0.965<sup>+</sup> | 0.052 |
|  | <u>Stance Time (s)</u> | 0.715<br>(0.061)* | 0.963<sup>+</sup> | 0.033 | 0.882 (0.154) | 0.983<sup>+</sup> | 0.056 | 0.974 (0.281) | 0.946<sup>+</sup> | 0.184 |
|  | <u>Swing Time (s)</u> | 0.363<br>(0.023)* | 0.913<sup>+</sup> | 0.020 | 0.401 (0.041) | 0.907<sup>+</sup> | 0.035 | 0.452 (0.097) | 0.964<sup>+</sup> | 0.051 |
| <u>Non paretic/Dominant</u> | <u>Step Length (m)</u> | 0.631<br>(0.062)* | 0.967<sup>+</sup> | 0.032 | 0.507 (0.102) | 0.978<sup>+</sup> | 0.042 | 0.455 (0.143) | 0.966<sup>+</sup> | 0.073 |
|  | <u>Stance Time (s)</u> | 0.717<br>(0.061)* | 0.967<sup>+</sup> | 0.031 | 0.883 (0.156) | 0.977<sup>+</sup> | 0.066 | 1.053 (0.317) | 0.977<sup>+</sup> | 0.135 |
|  | <u>Swing Time (s)</u> | 0.363<br>(0.024)* | 0.936<sup>+</sup> | 0.017 | 0.400 (0.040) | 0.963<sup>+</sup> | 0.022 | 0.373 (0.054) | 0.946<sup>+</sup> | 0.035 |

\* $p < 0.001$ compared to stroke
<sup>†</sup> $p < 0.001$ ICC significantly different from zero

### Reliability of Spatiotemporal Measures

Descriptive statistics for spatiotemporal parameters are presented in Table 2 for control participants (at SSS and Matched speeds) and participants post-stroke (at SSS). Most spatiotemporal parameters demonstrated excellent test–retest reliability (ICCs ranged from [0.873, 0.986]) in both groups and across walking conditions (Table 2). Step width was the only parameter with good reliability.

MDC values increased in control participants when walking at Matched (slower) speeds compared to SSS for all variables except step width (Table 2). When comparing post-stroke participants to controls walking at Matched speeds, MDC values were generally greater in participants post-stroke across most parameters, except stride length. MDCs for step lengths were greater for the non-paretic extremity, whereas MDCs for stance and swing times were greater for the paretic extremity.

### Reliability of Joint Range of Motion

Table 3 presents descriptive statistics for ROMs in the sagittal plane for control participants (at SSS and Matched speeds) and post-stroke participants (at SSS). At SSS, ROMs were significantly different for the paretic extremity compared to controls for the hip and knee (p=0.04 and p<0.001, respectively). At Matched speeds, only the paretic knee ROM was significantly different from controls (p=0.002). ICCs for ROMs ranged from poor to excellent reliability ([0.362, 0.960]). In controls, the hip showed good reliability at SSS and excellent reliability at Matched speed; the knee showed moderate reliability at SSS and moderate to good reliability at Matched speed; the ankle showed moderate reliability at SSS and Matched speeds. In participants post-stroke, the paretic extremity showed good reliability at the hip, excellent reliability at the knee and moderate reliability at the ankle. In contrast, the non-paretic extremity showed poor reliability at the hip, knee, and ankle, with all ICCs <0.50.

**Table 3:** Mean (Standard Deviation), ICC, and MDC values for range of motion for control participants walking at self-selected speed, controls walking at matched speed, and stroke participants.

|  | Variable | Control SSS |  |  | Control Matched |  |  | Stroke SSS |  |  |
| --- | --- | --- | --- | --- | --- | --- | --- | --- | --- | --- |
|  |  | Mean (SD) | ICC | MDC | Mean (SD) | ICC | MDC | Mean (SD) | ICC | MDC |
| Paretic/Non Dominant | Hip (degrees) | 43.923*<br>(5.280) | 0.870<sup>+</sup> | 5.392 | 38.812<br>(7.074) | 0.928<sup>+</sup> | 5.370 | 36.164<br>(9.296) | 0.815<sup>+</sup> | 11.263 |
|  | Knee degrees | 63.593*<br>(6.321) | 0.526 | 12.256 | 60.671*<br>(8.100) | 0.690<sup>+</sup> | 12.642 | 38.501 (18.612) | 0.960<sup>+</sup> | 10.531 |
|  | Ankle degrees | 23.717<br>(5.328) | 0.652 | 8.850 | 20.200 (4.297) | 0.518 | 8.418 | 19.622 (6.274) | 0.731<sup>+</sup> | 9.128 |
| Non paretic/Dominant | Hip degrees | 43.045<br>(5.339) | 0.859<sup>+</sup> | 5.670 | 37.233<br>(7.078) | 0.956<sup>+</sup> | 4.211 | 40.762<br>(6.382) | 0.362 | 14.355 |
|  | Knee degrees | 64.378<br>(7.847) | 0.640 | 13.213 | 60.629<br>(9.716) | 0.780<sup>+</sup> | 12.698 | 58.886 (8.903) | 0.498 | 17.786 |
|  | Ankle degrees | 24.229<br>(5.933) | 0.780<sup>+</sup> | 7.881 | 20.975 (4.828) | 0.738<sup>+</sup> | 6.990 | 23.176 (5.811) | 0.432 | 12.303 |

\*p significantly different compared to stroke
<sup>+</sup>p<0.001 ICC significantly different from zero

MDC values for ROM in control participants did not systematically change across speeds (Table 3). For participants post-stroke, MDC values were greater in the non-paretic compared to the paretic extremity for all joints. Additionally, compared to controls, MDC values in the paretic and non-paretic extremities were a greater percentage of the overall ROM (Figure 1); for example, for the paretic and non-paretic hip, the MDC in stroke was ~30% of the overall ROM, for the knee it was ~25% of the overall ROM, and for the ankle it was ~50% of the overall ROM (compared to ~13%, 20% and 35% for the control hip, knee and ankle, respectively).

**Figure 1.**
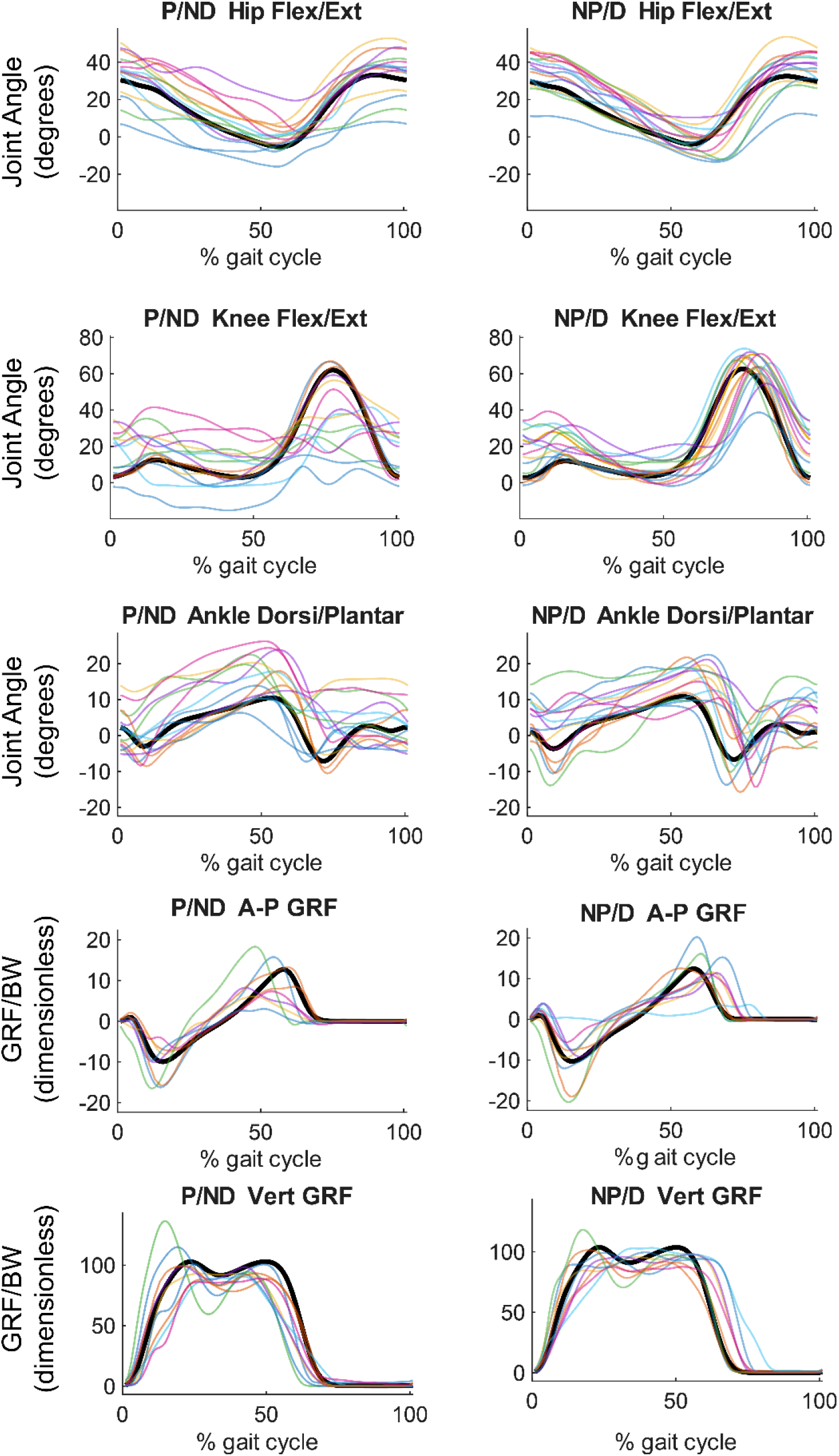
Joint kinematics and GRFs over the gait cycle. Timeseries data are presented for all stroke participants in colored lines, relative to the average of control participants during the Matched conditions in bold. P: Paretic. ND: Non-dominant. NP: Non-paretic. D: Dominant. GRF: Ground reaction force. BW: Body Weight. A-P: Anterior-Posterior. Vert: Vertical.

### Reliability of ground reaction forces

Table 4 presents descriptive statistics for peak propulsive and vertical GRFs for control participants (at SSS and Matched speeds) and post-stroke participants (at SSS). Peak propulsive GRFs were greater in controls at SSS compared to participants post-stroke (p<0.01). Peak propulsive GRFs did not differ at Matched speeds. ICCs for peak propulsive GRFs showed excellent reliability both for control and stroke participants. ICCs for peak vertical GRF showed good reliability in controls and excellent reliability post-stroke.

**Table 4:** Mean (Standard Deviation), ICC, and MDC values for peak propulsive force and peak vertical ground reaction force.

|  | Variable | <u>Control SSS</u> |  |  | <u>Control Matched</u> |  |  | <u>Stroke SSS</u> |  |  |
| --- | --- | --- | --- | --- | --- | --- | --- | --- | --- | --- |
|  |  | <u>Mean (SD)</u> | <u>ICC</u> | <u>MDC</u> | <u>Mean (SD)</u> | <u>ICC</u> | <u>MDC</u> | <u>Mean (SD)</u> | <u>ICC</u> | <u>MDC</u> |
| <u>Paretic/Non Dominant</u> | <u>Peak pGRF (GRF/BW)</u> | 0.185 (0.038)* | 0.990<sup>+</sup> | 0.011 | 0.130 (0.045) | 0.994<sup>+</sup> | 0.010 | 0.095 (0.058) | 0.983<sup>+</sup> | 0.022 |
|  | <u>Peak vGRF (GRF/BW)</u> | 1.137 (0.067)* | 0.953<sup>+</sup> | 0.042 | 1.072 (0.035) | 0.783<sup>+</sup> | 0.047 | 1.034 (0.147) | 0.982<sup>+</sup> | 0.056 |
| <u>Non paretic/Dominant</u> | <u>Peak pGRF (GRF/BW)</u> | 0.180 (0.040)* | 0.987<sup>+</sup> | 0.013 | 0.129 (0.045) | 0.983<sup>+</sup> | 0.017 | 0.127 (0.050) | 0.982<sup>+</sup> | 0.019 |
|  | <u>Peak vGRF (GRF/BW)</u> | 1.140 (0.062)* | 0.870<sup>+</sup> | 0.064 | 1.077 (0.024)* | 0.778<sup>+</sup> | 0.032 | 1.013 (0.066) | 0.912<sup>+</sup> | 0.057 |

\*p significantly different compared to stroke
<sup>+</sup>p<0.001 ICC significantly different from zero
pGRF: peak propulsive ground reaction force
vGRF: peak vertical ground reaction force
BW: body weight

In post-stroke participants, MDCs were greater than in controls at matched speeds for both propulsive and vertical GRFs. The MDCs in people post-stroke also represented a greater percentage of overall propulsive and vertical GRF. Particularly for the propulsive GRF, the MDC was 23% of the total propulsive GRF for the paretic extremity and 15% for the non-paretic extremity compared to 13% in controls.

## Discussion

The primary objective of this study was to quantify test-retest reliability ICC and MDC values for gait analysis outcomes in individuals post-stroke and age-, sex-, and speed-matched neurotypical controls. By including a matched-speed control condition, we assessed whether post-stroke impairment increases variability and reduces gait analysis reliability, or whether post-stroke impairment alone accounts for post-stroke gait analysis reliability. Overall, contrary to our hypothesis, we found that ICCs were not systematically lower in post-stroke participants; some measures showed greater reliability post-stroke than in controls, indicating more replicated gait behaviors between days post-stroke. Additionally, although some post-stroke variables had higher ICC than controls, almost all variables showed greater MDC values in post-stroke participants than in speed-matched controls, consistent with our hypothesis. We found that: 1) spatiotemporal variables showed comparable ICCs between stroke and control, yet MDC values remained greater in the post-stroke group even when walking speed and spatiotemporal values were matched to controls. 2) ROM ICCs were greater for the paretic extremity, yet MDCs represented a larger percentage of overall ROM post-stroke compared to controls. We also found that MDCs in the non-paretic extremity were greater compared to the paretic extremity. 3) GRFs show excellent reliability post-stroke, yet post-stroke MDCs for peak propulsive and vertical GRFs are greater than in controls. Together, these findings indicate that both paretic impairments and non-paretic compensations contribute to measurement reliability and influence the effect size of interventions needed to ensure measured changes are beyond measurement error in post-stroke populations.

Our analyses further highlight the challenges of joint kinematics as outcome measures in post-stroke rehabilitation studies. Surprisingly, we found that ICCs for the paretic extremity were not only greater compared to the non-paretic extremity, but they were also greater than in controls. In addition, MDC values were consistently larger in the non-paretic than in the paretic lower extremity. The combination of these results points to motor constraints in the paretic lower extremity (Sánchez et al., 2018; Sanchez and Dewald, 2014) that may reduce variability during walking from day to day, and compensatory patterns in the non-paretic extremity (Chen et al., 2005; Kuch et al., 2025; Raja et al., 2012) that may increase variability from day to day. These findings indicate different mechanisms for variability in the paretic compared to the non-paretic extremity and compared to controls, consistent with the idea of reduced flexibility during gait post-stroke (Clark et al., 2010), which directly influences the reliability of gait analysis. These results also show that large changes in joint kinematics are required before they can be confidently interpreted as true biomechanical improvements.

The ICC and MDC values obtained here are consistent with previous studies examining gait reliability in post-stroke populations during both treadmill (Kesar et al., 2011) and overground walking (Geiger et al., 2019). For treadmill walking, Kesar et al. reported for paretic step length an MDC of 0.0675 m. In comparison, the present study observed a smaller MDC (0.052 m), suggesting a lower threshold for detecting meaningful change, which can be explained by our sample of participants having less impairment (average FMA 23.1 compared to 19 in Kesar). For the nonparetic extremity, we observed a larger MDC (0.073 m in our study vs. 0.056 m), indicating somewhat greater variability on the non-paretic side, contrary to prior studies (Balasubramanian et al., 2009). The Kesar study also reported MDCs for peak propulsive and vertical GRFs; MDC values in our study for the paretic propulsive GRF were about twice those reported in this previous study. Similarly, Kesar et al. reported the vertical GRF averaged across limbs with an MDC smaller than our study. Comparison with overground walking data (for the paretic extremity only) as reported by (Geiger et al., 2019) shows strong agreement in reliability estimates across most spatiotemporal variables. ICC values for step width were lower, pointing to potential increased variability in step widths during treadmill walking compared to overground walking (Rosenblatt and Grabiner, 2010). MDC estimates followed a similar pattern: we observed smaller MDCs for stride length (0.078 m in our study vs. 0.1273 m), cadence (10.47 in our study vs. 11.81 steps/min), and paretic step length (0.052 m in our study vs. 0.0702 m), but larger MDCs for nonparetic step length and step width. Geiger et al., also reported ROM values for the paretic extremity. Overall, our study observed a greater paretic ROM for the hip and knee (36 and 38 degrees, respectively) compared to Geiger (25 and 23 degrees), and consistently, greater MDCs (~11 degrees in our study for the hip and knee compared to ~5 and 8 degrees in Geiger). These differences between studies likely reflect task-specific factors such as treadmill vs. overground walking and differences in walking speed and impairment between study samples (walking speeds were not reported for the Geiger study).

To contextualize the clinical relevance of the present MDC values, we compared the MDC obtained here with changes reported in intervention studies post-stroke. In our previous work (Sánchez and Finley, 2018), participants used targeted visual feedback to reduce step length asymmetry leading to increases in non-paretic step length of 53 mm or increases in paretic step length of 19 mm, both of which are below our MDCs. Using split-belt walking, (Reisman et al., 2013) reported changes in step length of ~0.06 m for the paretic and ~0.09 m for the non-paretic extremity, which exceed the corresponding MDC identified here, while increases in cadence of approximately 9 steps/min fall below the MDC of our study (11.47 steps/min). Using a user-driven treadmill, (Donlin et al., 2021) reported reductions in step width ~0.031 m that did not exceed the MDC identified here (0.065 m). A recent high-intensity locomotor training study (Ardestani et al., 2020) showed changes in cadence of 23 steps/min and changes in stride length of 0.12–0.15 m, which exceeded MDC thresholds. Similarly, studies targeting paretic propulsion showed group-level increases in propulsive force of around 4% body weight using electrical stimulation (Choe et al., 2024), ~11% body weight using exoskeleton assistance (Awad et al., 2017), and ~4% body weight using propulsion biofeedback (Hinton et al., 2024; Santucci et al., 2023), all exceeding the ~2% MDC value from our study. In contrast, a study using kinematic feedback of hip extension only saw increases in trailing limb angle of ~3 degrees (Hinton et al., 2024), which, while significant, are well below MDC values of 11 degrees observed in our study. Collectively, these comparisons demonstrate that while all interventions observed significant differences due to the interventions, only some, but not all, previous interventions achieved changes above the margin of measurement error in people post-stroke.

### Limitations

This study has several limitations. First, the post-stroke sample was limited to individuals with mild-to-moderate impairments; therefore, the present MDC values may underestimate variability in more severely impaired populations. Second, although control participants were matched by age and sex, other factors that may influence gait variability, such as body mass index, which influences identification of bony landmarks for marker placement (Wolf et al., 2009), were not controlled or used for matching. Third, clothing was not standardized across participants, which may have introduced variability in marker tracking due to marker occlusion or displacement. In addition, all data were collected during treadmill walking, which differs from overground locomotion and may limit generalizability to real-world conditions. Finally, our analyses were limited to sagittal plane ROMs and peak GRFs. Future studies should examine MDC values in all three planes for kinematics and kinetics.

## Conclusion

This study quantified test-retest reliability ICC and MDC for key spatiotemporal, kinematic, and kinetic parameters in individuals post-stroke and matched neurotypical controls. By incorporating a speed-matched control condition, we showed that increased post-stroke MDC values reflect stroke-related gait variability beyond the effects of walking speed or methodological limitations alone; differences in MDCs between the paretic and non-paretic extremities also point to mechanistic differences that influence variability and thus reliability between extremities. Our findings provide important benchmarks for interpreting longitudinal changes in gait in individuals post-stroke and also in neurotypical controls.

## Data Availability

All data produced in the present study are available upon reasonable request to the authors

## Acknowledgements

This research was funded by the National Center for Medical Rehabilitation Research grant R03HD107630 and the National Center for Advancing Translational Sciences grant R03TR004248, both to Natalia Sánchez.

## Authors contribution

AAR: Investigation, Methodology, Data curation, Formal analysis, Software, Writing – original draft, AK: Investigation, Data curation, Formal analysis, Software, Visualization, Writing – review & editing, RTJ: Conceptualization, Writing – review & editing, NS: Conceptualization, Funding acquisition, Methodology, Supervision, Writing – original draft, Writing – review & editing.

